# Effect of food-cues on aMCC glutamate modulation during inhibitory control post weight loss intervention: a ^1^H fMRS study

**DOI:** 10.64898/2026.10.27.26366634

**Authors:** Jillian M. Eichstaedt, Rachel Sochocki, Dalal Khatib, Nicole Miller, Vaibhav A. Diwadkar, Paul Burghardt, Amy Rothberg, Jeffrey A. Stanley

## Abstract

**Objective:** Poor inhibitory control may decrease weight loss maintenance. The neurobiological mechanisms are poorly understood. We investigated whether glutamate modulation in the anterior mid-cingulate cortex (aMCC) during response inhibition is impacted by food-cues and associated with self-reported behaviors and weight loss maintenance.

**Methods:** 27 adults with overweight/obesity were recruited from a clinical weight management program. ^1^H fMRS (midline aMCC) was acquired during a visual-guided response inhibition task engaging two responding modes [non-selective (100% responses) or selective (withholding a response on 20% of trials)] applied with neutral (squares) or food/non-food cues. The impact of stimulus type on the aMCC glutamate modulation for each responding mode was assessed using a repeated measures GEE approach. Correlations were conducted between glutamate modulation, Three-Factor Eating Questionnaire scores, and long-term weight change.

**Results:** The stimulus effect was significant during selective responding, driven by lower aMCC glutamate modulation during food/non-food cues. This glutamate modulation (selective; food/non-food) association with weight loss maintenance was a close trend (*R^2^*=0.15, *p*=0.054). The association with the TFEQ Cognitive Restraint was significant (*R^2^*=0.20, *p*=0.047).

**Conclusions:** These results demonstrate an impact of food/non-food cues during response inhibition on the neurobiology and associations with long-term weight loss maintenance and scores related to eating behaviors.

## Introduction

Obesity is a highly prevalent condition with increased risk of health-related comorbid conditions such as heart disease, type 2 diabetes, and multiple cancers, which are preventable with proper treatment [1,2]. Though weight reduction is a core objective in obesity treatment, it is estimated that following weight loss, most individuals will regain 30–80% of lost weight within 5 years [3]. While the physiology changes associated with reducing weight have been well categorized [3], full understanding of why some individuals cannot maintain a reduced weight remains unclear. This stresses the importance of identifying cognitive/behavioral and neurobiological markers that can predict one’s ability to maintain long-term weight loss and lead to the development of better targeted interventions.

There is increasing evidence associating poor top-down inhibitory control with the inability of maintaining long-term weight loss in individuals with obesity [4]. Inhibitory control can be conceptualized as the ability to suppress certain thoughts, actions, or, in the context to obesity, urges to consume excess food [4,5]. Inhibitory control can be assessed multiple ways, including using the Go/No-Go, Stop-Signal or the Delayed Discounting task [5]. The Go/No-Go, among the most commonly used tasks, is typically conducted as a visual guided motor response inhibition task with rapidly presented cues [i.e., neutral objects or alphanumeric digit(s)] where motor responses are selectively withheld on infrequent “no-go” trials (or target cues) while motor responses are required on frequent “go” trials (or non-target cues) [5,6]. A recent meta-analysis summarized that studies applying the Go/No-Go task have reported poorer performance accuracy in individuals with obesity compared to normal weight [7]. Go/No-Go task difficulty can be modified by swapping neutral objects/digits with cues containing appetitive food item images, which may interfere with the top-down control of inhibitory processes by introducing competing thoughts such as urges of consuming excess foods [4,8]. It has been identified that food-based cues/stimuli, but not neutral cues, impair inhibitory control performance in individuals with obesity [7]. It is presumed that an individual with good top-down inhibitory control ability would not be influenced, or be distracted, by the presence of appetitive food cues. However, the association between food cues impairing inhibitory control and the ability to maintain long-term weight loss in obesity is unclear.

The anterior mid-cingulate cortex (aMCC, also commonly referred to as the dorsal anterior cingulate cortex, dACC) is a central brain area for top-down inhibitory control processing. This includes the monitoring and differentiating of stimuli for selective inhibitory motor responses and moderating motor responses to visual stimuli in coordination with the supplementary motor area [5,9–11]. Given the noted deficits, task-based fMRI studies have implicated the aMCC during inhibitory control demand in obesity, demonstrating a lack of increased BOLD activation in the aMCC of individuals with obesity compared to non-overweight individuals [12,13]. Reduced BOLD activation in the ACC during a Stop-Signal task (contrasting error trials with correct stop trials using neutral cues) has further been associated with poorer ability in losing body fat over a 6-month period in adults with overweight/obesity (OV/OB) [14]. However, the extent of the interpretation of a reduced activation and its association with weight loss ability is limited due to the BOLD fMRI signal being driven by the hemodynamic response of brain tissue, which is viewed as a distal proxy reflecting neural activity or processes [15,16].

Neural activity is driven by the neurotransmission of the well-integrated excitatory and inhibitory neurotransmitters, glutamate and gamma-aminobutyric acid (GABA), respectively [17–19]. ^1^H functional magnetic resonance spectroscopy (^1^H fMRS) can detect temporal changes in the steady-state level of glutamate (and/or GABA) under different task conditions or demand [20–23]. Previous ¹H fMRS work by our group has demonstrated significantly different levels of increased glutamate modulation in the aMCC during “all-go” (or non-selective reflecting generalized processes for motor control) and “go/no-go” (or selective reflecting processes of response inhibition plus generalized motor control) responding modes compared to each other or the baseline control condition (fixation crosshair) utilizing neutral cues in healthy adolescents, suggesting a potential differential increased excitatory neuromodulation drive in the aMCC during inhibitory control processes [24]. This provides a framework for investigating the neurobiology related to differences in the aMCC glutamate modulation during response inhibition performance (non-selective and selective responding modes) with or without appetitive food cues.

The above supports poorer inhibitory control performance in the presence of appetitive food cues in individuals with obesity and reduced fMRI BOLD activation in the aMCC under inhibitory control; however, there is little-to-no evidence supporting behavioral/neurobiological measurements that can predict one’s ability of maintaining long-term weight loss. This study is leveraging a unique sample of adults with OV/OB who participated in the Michigan Weight Management Program (MWMP - an intensive lifestyle intervention that targets a 15% body weight reduction at 3 months and long-term weight reduction [25]) of ≥ 10% 6-months, 12- and 24-months after starting the intervention (reflecting weight stabilization). The ability to maintain long-term weight loss was assessed at 12-months. At 6-months (prior to the maintenance period), ¹H fMRS from the aMCC utilizing a response inhibition task (non-selective and selective responding modes) with and without food cues and eating behavior metrics using the Three Factor Eating Questionnaire (TFEQ [26]). This allowed us to investigate in individuals with OV/OB: 1) the impact aMCC glutamate modulation during inhibitory control performance on the presence of food-cues compared to neutral cues; 2) whether higher glutamate modulation during inhibitory control with food-cues, reflecting a lesser impact on aMCC glutamate when presented with food-cues, can predict greater ability to maintain long-term weight loss during the maintenance period (6-to 12-months); and 3) whether higher TFEQ restraint scores are associated with higher aMCC glutamate modulation with food-cues and predict greater ability in maintaining long-term weight loss.

## Methods

### Participants

A total sample of 27 adults with OV/OB who were enrolled in the MWMP were recruited for this study. The MWMP is a two-year long, multicomponent, structured, clinical research weight-loss intervention program [25] that employs an intensive caloric restriction for the first 12 weeks to promote 15% weight loss followed by regular physician and dietitian interaction to support change in diet and physical activity behaviors for long-term weight control. Study inclusion criteria included being between 20 and 65 years of age, having recently completed the first 6 months of the intervention with a weight reduction from baseline body weight of at least ≥10% (a percent body weight reduction associated with improvements in cardiometabolic outcomes [27]), and being able and willing to provide written informed consent for study participation. The 6-month time-point was the point when the focus shifted from weight loss to weight loss maintenance. Exclusion criteria for the MWMP, which extended to this study, included a starting BMI at program entry <30, previous bariatric surgery, a weight loss of >5 kg prior to program enrollment, current self-reported cigarette smoking, and past/present diagnosis of a substance use/dependent disorder, major psychiatric disorder or eating disorder, and current or planned pregnancy within one year. Additional exclusion criteria for this study included MRI contraindications (including a weight of ≥158 kg). All participants provided informed consent for study participation, and this study was approved by the Institutional Review Board at Wayne State University (IRB-21-01-3088).

### Weight and Self Report Questionnaire

Participant weight data was collected by the MWMP, which included weight collected at enrollment (prior to weight loss), 6-month time-point (end of the weight stabilization phase) and 12-month time-point (reflecting the weight following 6 months of maintenance). Additionally, eating behavior measurements (TFEQ [26] scores) were collected at the 6-month time-point. The domain of interest is Cognitive Restraint, which reflects the confidence in suppressing urges to eat, particularly in the absence of hunger cues and conceptually aligns with inhibitory control ability. The other two domains include Emotional Eating (reflecting likelihood to eat under significant emotional distress) and Unrestrained Eating (reflecting likelihood to eat large quantities of food at a time with a sense of feeling out of control).

### Motor Response Inhibition Task

This study utilized a visually-guided motor response inhibition task adapted from fMRI [9] that included four separate task runs; one for each motor responding mode (non-selective and selective) and in each utilizing neutral cues (red/green squares) and food/non-food cues (appetitive food/neutral non-food objects) (**Figure 1**). The non-selective (‘all-go’) responding mode included flashing probes (either red/green squares or food/non-food cues) presented in a randomized 50:50 (red:green or food:non-food) distribution. Participants were instructed to respond as quickly as possible to all cues (100% motor responses; 0% inhibition). During the selective (‘go/no-go’) responding mode, participants were instructed to respond as quickly and accurately as possible in response to only ‘go’ cues (either green squares or food objects; 80% of the presented cues to ensure prepotent inhibition response) and to selectively inhibit responses to ‘no-go’ cues (either red squares or non-food objects; 20% of the presented cues). For all motor responses, participants were instructed to tap their forefinger on the surface of a touchpad (fiber-optic response pad by Current Design Systems, Inc.). The food/non-food objects were selected from the “Food-Pics Extended” database, which included highly palatable and recognizable food images and non-food images with neutral arousal and valance [28]. All cues were presented for 0.25s with a randomly jittered interstimulus intervals between 1.0 – 1.5s in length. Each task run consisted of four repeated 32s epochs and each separated by a 16s of passive visual fixation of a static crosshair **(Figure 1)**. The anticipated sample size was projected to be inadequate for randomized sampling and therefore, the order of the task runs was fixed with the non-selective responding mode always ahead of the selective responding mode. Further, the two responding modes with neutral cues were always conducted before the two responding modes with food/non-food cues. A baseline comparison condition (four repeated 32s epochs of visualizing a flashing checkerboard interspersed with 16s rest epochs) was acquired as a separate run preceding the task conditions. The tasks (programmed using Presentation; Neurobehavioral Systems Inc.) were projected onto a screen positioned at the rear of the scanner for participants to view with an angled mirror attached to the head coil. Moreover, the motor response performance data was collected to estimate the mean response latency, hit rate, and false alarm rate (selective mode only) for each epoch within the task runs.

**Figure 1.**
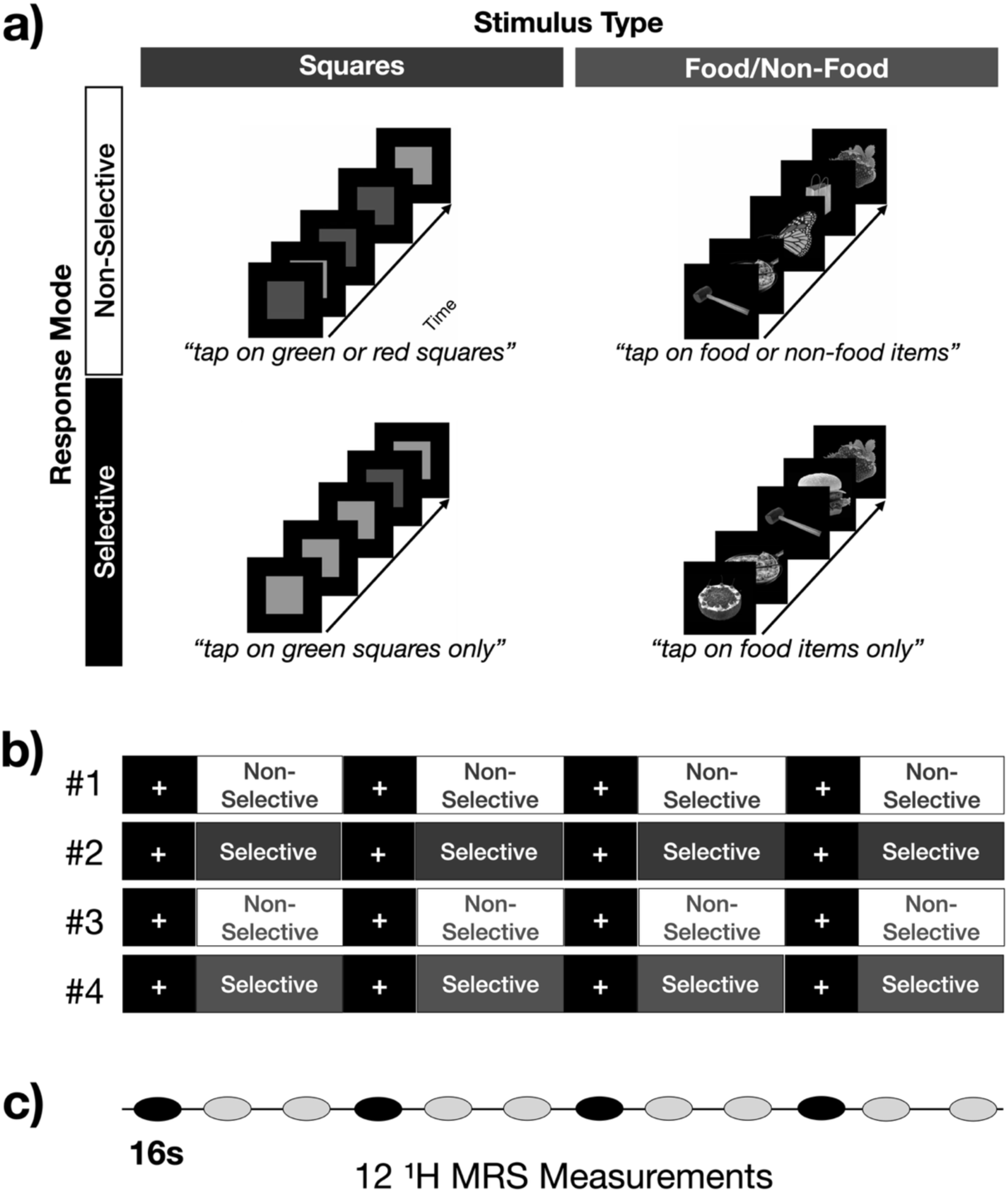
^1^H fMRS inhibitory motor control task with neutral and food/non-food stimuli. (a) The two responding modes (non-selective and selective) and two stimulus types [neutral squares (light grey reflecting green squares; darker grey reflecting red squares) and food/non-food objects) and shown. (b) The chronological order of the four task runs each contained four 32s epochs of task separated by 16s rest blocks of passive visual fixation. (c) Twelve 16s ^1^H fMRS measurements were collected during each task run. The two ¹H fMRS measurements in each task epoch were averaged to yield four datapoints per task run per participant to be statistically analyzed.

**Figure 2.**
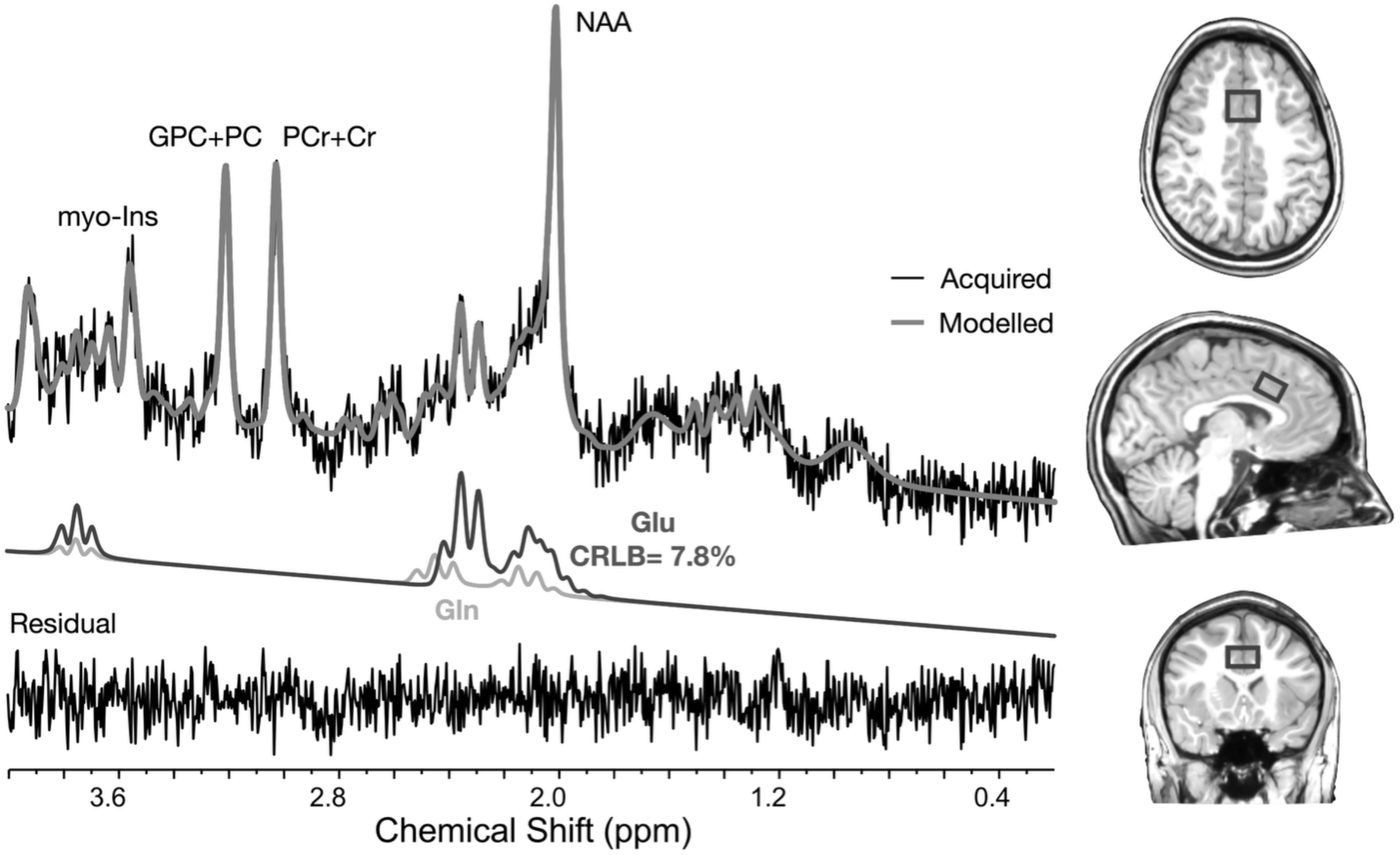
An example of an averaged ^1^H fMRS spectrum composed of 2 16s spectra (32s temporal resolution). Data shown include the fitted signal, glutamate (Glu), and glutamine (Gln). The aMCC voxel placement is shown on the structural images by a box. Abbreviations: myo-Inositol (myo-Ins); glycerophophocholine plus phosphocholine (GPC+PC); phosphocreatine plus creatine (PCr+Cr); N-acetyl aspartate (NAA); glutamine (Gln); and glutamate (Glu).

### ^1^H fMRS

^1^H fMRS data was collected on a 3T Siemens system (Verio or Cima.X) system with a 32-channel head coil. All scans were conducted in the morning to control for possible diurnal effects and standardize time between meals. First, T_1_-weighted structural images were collected (MPRAGE; TR=2150ms; TE=3.2ms; TI=1100ms; flip-angle=8^0^; FOV=256x256x160 mm^3^; 160 axial slices of thickness=1mm; pixel resolution 1x1x1 mm^3^; acquisition time=4:59min) and subsequently used for placement of the ^1^H fMRS voxel. Placement of the aMCC voxel location and orientation utilized the auto-voxel placement procedure to co-register the participant T_1_-weighted image to the predefined template voxel, ensuring consistency in voxel placement across participants [29]. The ^1^H fMRS data was acquired utilizing a PRESS sequence (developed by Edward J. Auerbach and Małgorzata Marjańska and provided under a C2P agreement by the University of Minnesota), which included outer volume saturation (OVS), variable power and optimized relaxation times (VAPOR) for water suppression [30], and the following parameters: TR=4.0s, TE=23ms, 2048 complex data points; 2 kHz bandwidth; 12 measurements per task run; 4 averages/measurement (16s temporal resolution), acquisition time=3:19min per task run. Shimming over the aMCC voxel area to optimize the spectral linewidths utilized FAST(EST)MAP [31]. A fully relaxed unsuppressed water measurement was collected following each task run for spectral correction and absolute quantification (TR=10s, 2 averages, acquisition time=0:20min) [32–34].

The first measurement of each task run was removed for post-processing due to the potential of the signal not reaching steady-state equilibrium. The data collected during the 16s fixation cross rest measurements was also excluded to prevent carryover effects. LCModel was utilized for spectral processing and quantification [35], in combination with in-house scripts and programs. For the remaining ¹H fMRS measurements, spectral phase (0^th^ and 1^st^-order) and frequency shift corrections were estimated using the using the LCModel LCORAW option. For each task run, an averaged 1^st^ order phase value from the ¹H fMRS measurements was calculated. All ¹H fMRS measurements within a task run were then re-corrected for phase and frequency shift (LCORAW option) by fixing the 1^st^ order phase with the averaged value, as previous described [36]. Following this correction, the two consecutive ^1^H fMRS measurements within a task epoch were averaged and fitted for analyses. The simulated basis set for LCModel fitting included N-acetyl aspartate (NAA), N-acetyl aspartyl glutamate (NAAG), phosphocreatine (PCr), creatine (Cr), glycerophosphocholine (GPC), phosphocholine (PC), myo-Inositol, glutamate, glutamine, GABA, lactate, glutathione (GSH), aspartate, alanine, glucose, taurine, scyllo-Inositol, and macromolecules/lipids. A soft constraint on the glutamate to glutamine ratio was also applied to ensure a reasonable physiological range [i.e., CHRATIO(14) was set to “Glu/Gln = 4.0002±2.6668” or a glutamate/glutamine ratio range between 6.6667 and 1.3334] [36–38]. The primary ^1^H MRS metabolite of interest was glutamate, but additional key metabolites – NAA, PCr+Cr, GPC+PC, and myo-Inositol – were also extracted for further analyses. The net result was one estimate of glutamate and other key metabolites per task epoch, or four distinct metabolite values per response mode/stimulus type. Any spectral results with a SNR<5 or CRLB>20, or an epoch condition with only a single non-excluded measurement, were deemed as poor quality and excluded from the analyses. Tissue segmentation analyses were performed on the T_1_-weighted images using FSL ANAT, and the estimated tissue fraction values within each voxel along with the unsuppressed water signal and relaxation values were utilized for absolute quantification [33,34].

## Statistical Analyses

Repeated measures generalized estimating equation (rmGEE; SAS GENMOD, SAS Institute Inc) analyses were conducted to address the impact of stimulus type (neutral squares vs. food/non-food objects) on aMCC glutamate modulation during inhibitory control performance of the motor responding modes, non-selective and selective. Two models were conducted, one per responding mode (non-selective and selective). The dependent variable was the absolute glutamate level, and the model terms were stimulus type (neutral squares or food/non-food objects), glutamate level from the baseline condition, age, and sex. A similar approach utilizing rmGEE was taken to assess the behavioral performance data for each motor responding mode separately, with response latency, hit rate, and false alarm rate (selective mode only) as the dependent variable. Stimulus type, glutamate modulation (percent change relative to the baseline comparison condition), interaction between stimulus type and glutamate modulation, age, and sex were the independent variables. Correlation analyses were conducted between the aMCC glutamate modulation (specifically during processes related to selectively inhibiting motor responses in the presence of appetitive food/non-food cues) and weight loss ability during the long-term maintenance period (12-month minus 6-month) and self-reported eating behaviors (TFEQ Cognitive Restraint scores) to assess their relationship. A Bonferroni correction for multiple comparisons was not applied due to the small sample size. Therefore, the threshold for statistical significance was *p* ≤ 0.05.

## Results

### Sample and ^1^H fMRS Data Quality

Of the 27 participants, 22 were scanned at the 6-month time point, whereas the remaining 5 participants were scanned between 10 and 13 months following the MWMP entry time due to logistical issues. Participant demographics are shown in **Table 1**. The overall quality of the ^1^H fMRS spectra was high, exhibiting a mean (±SD) sign-to-noise ratio (SNR) of NAA of 7.8±1.6, full-width-at-half-max (FWHM) of 4.9±1.2 Hz, and Cramer-Rao lowest bound for glutamate (CRLB) of 8.0±1.4, as shown in **Table 2**. A total of 34 ¹H fMRS spectra were excluded due to not meeting the data quality criteria, which reflected a 7.9% attrition rate. The mean grey matter tissue fraction (±SD) within the aMCC voxel was 47.3±6.3% (**Table 2**). The mean (±SD) overlap of the ¹H fMRS voxel placement across all participants when transformed to the standard MNI space was 70.4±10.3%. There was no difference in the FWHM values during task active conditions compared to the control condition (*χ^2^* =5.30, *p*=0.26, see **Table S1**).

**Table 1.** Participant demographics and weight loss performance.

| **Table 1.** Participant demographics and weight loss performance. |  |  |
| --- | --- | --- |
| **Sample Size** | 27 |  |
| **Sex Distribution** | 15 Females and 12 Males |  |
|  | **Mean (±SD)** | **Min/Max Range** |
| **Age in years** | 50.3 (8.7) | 32.0 to 65.8 |
| **BMI at MWMP entry** | 39.5 (5.5) | 31.6 to 49.4 |
| **Weight at MWMP entry in kg.** | 118.2 (21.8) | 80.8 to 158.3 |
| **% Weight Change** |  |  |
| **6-month minus MWMP entry** | -20.8 (5.7) | -35.6 to -12.3 |
| **12-month minus 6-month** | 1.4 (4.6) | -8.6 to 8.4 |

**Table 2.** Mean absolute metabolite levels (institutional units) and spectral quality metrics of aMCC ¹H MRS data across all conditions.

| **Metabolites** | **Absolute Level** | **% CRLB** |  |
| --- | --- | --- | --- |
|  | In institutional units |  |  |
|  | **Mean (±SD)** | **Mean (±SD)** | **Min/Max Range** |
| Glutamate | 13.1 (2.1) | 8.0 (1.4) | 6 – 16 |
| NAA | 10.7 (1.5) | 5.2 (1.2) | 3 – 10 |
| PCr+Cr | 8.5 (1.2) | 4.8 (0.9) | 3 – 7 |
| GPC+PC | 2.5 (0.5) | 5.4 (1.1) | 3 – 11 |
| myo-Inositol | 8.2 (1.5) | 6.3 (1.4) | 4 – 18 |

| **Spectral Quality Metrics** | **Mean (±SD)** | **Min/Max Range** |
| --- | --- | --- |
| SNR NAA | 7.8 (1.6) | 5 – 11 |
| FHWM (Hz) | 4.9 (1.2) | 3.0 – 10.7 |

| **Tissue Fraction within the Voxel** | **Mean (±SD)** | **Min/Max Range** |
| --- | --- | --- |
| Grey Matter | 0.473 (0.063) | 0.366 – 0.620 |
| White Matter | 0.194 (0.051) | 0.057 – 0.279 |
| CSF | 0.333 (0.079) | 0.219 – 0.511 |

### ¹H fMRS Glutamate Modulation

Regarding differences in the aMCC glutamate modulation, the stimulus type term was not significant during the non-selective responding mode (*χ^2^* =0.95, *p*=0.33) but was significant during the selective mode (*χ^2^* =4.31, *p*=0.038) exemplified by lower aMCC glutamate modulation when selectively responding to food/non-food cues compared to neutral squares (**Figure 3**). Both the sex and age terms were not significant for the non-selective (*χ^2^* =0.71, *p*=0.40 and *χ^2^* =0.09, *p*=0.76, respectively) or selective (*χ^2^* =0.55, *p*=0.46 and *χ^2^* =2.21, *p*=0.14, respectively) responding modes.

**Figure 3.**
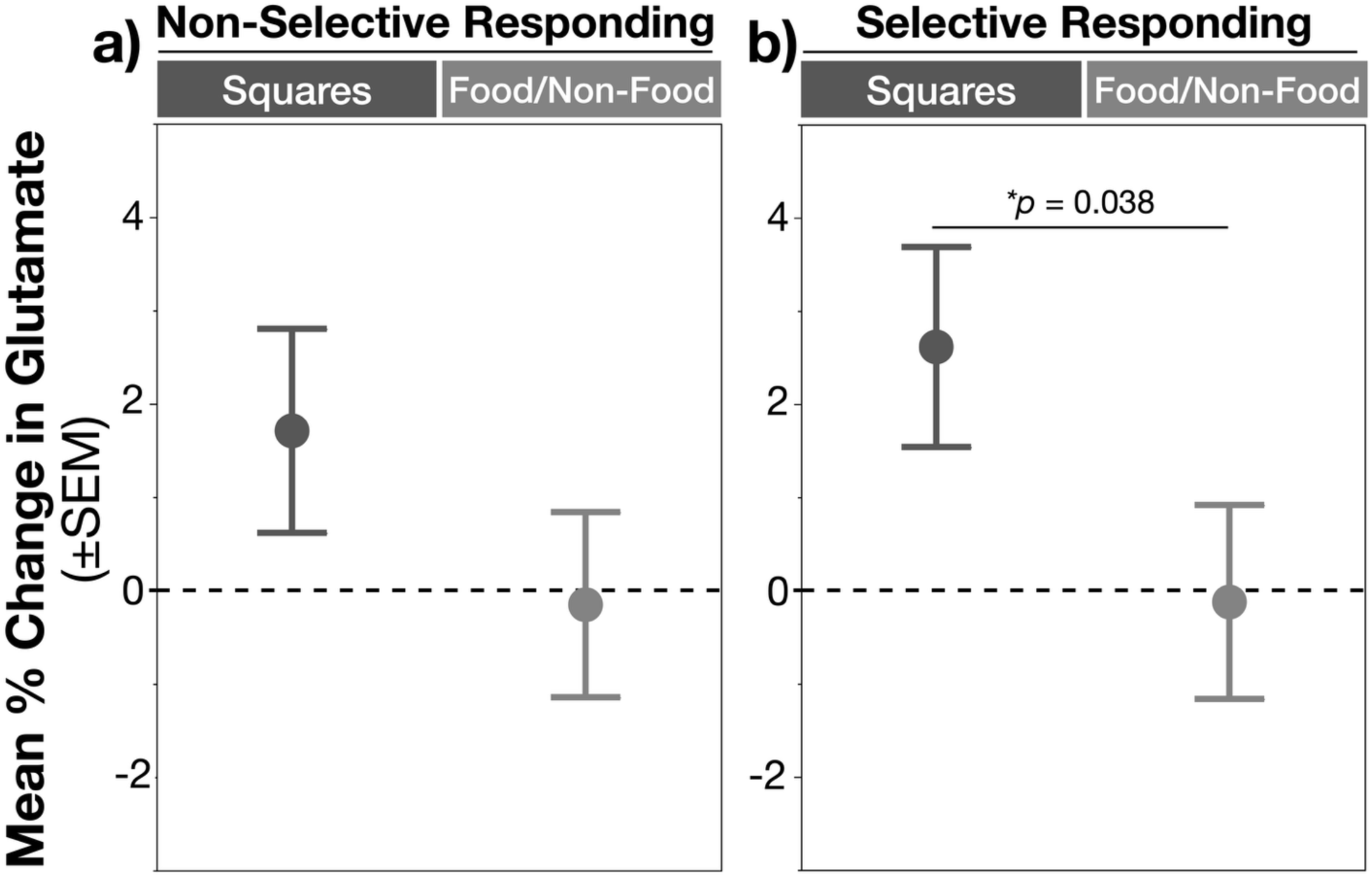
Mean glutamate modulation from the aMCC across Stimuli types for each Responding Mode expressed as percent change relative to the baseline comparison condition (fixation crosshair). No significant difference between stimulus types was observed during the non-selective responding mode (a), but a significantly higher aMCC glutamate modulation was observed during the selective mode in the presence of neural (squares) cues compared to food/non-food cues (b).

### Motor Response Inhibition Task Performance

Due to technological difficulties, tapping performance data was not available for one participant. Stimulus type was not significant during the non-selective or selective responding modes, for response latency or accuracy (or response hit rate); see **Table S2**. However, the stimulus type term was significant for false alarm rate, which is specific to the selective responding mode (*χ^2^* =5.14, *p*=0.023; **Table S2**). This led to significantly higher false alarm rates when presented with appetitive food/non-food cues compared to neutral (squares) cues **(Figure S1)**. The aMCC glutamate modulation term was not significant in the non-selective or selective responding modes for motor response latency (*χ^2^* =1.28, *p*=0.26 and *χ^2^* =2.17, *p*=0.14), response hit rate (*χ^2^* =0.02, *p*=0.89 and *χ^2^* =0.79, *p*=0.37), or false alarm rate (*χ^2^* =0.03, *p*=0.86), providing no evidence of behavioral motor tapping performance being moderated by the ¹H fMRS glutamate signal from the aMCC. **Table S2** includes a complete description of the results including the effects on Age and Sex.

### Associations between Neurobiology, Long-Term Weight Loss Ability and Eating Behaviors

The overall weight change during the long-term maintenance period (12-month minus 6-month) was 1.4±4.6%, which reflects an overall regain in weight but also a considerable range in weight loss/gain performance (-8.6%–8.4% across participants; **Table 1**). A series of correlations analyses were conducted to determine if a greater impact (reduction) in aMCC glutamate modulation during the selective responding mode during food/non-food cues (demonstrated to be significantly different) was associated with poorer ability to maintain long-term weight loss or with lower Cognitive Restraint scores (TFEQ). A close trend of lower aMCC glutamate modulation predicting poorer ability in the long-term maintenance of weight was observed [*R^2^*=0.15, *F*(1,23)=4.13, *p*=0.054, **Figure 4a].** Unfortunately, TFEQ scores were only available for 20 participants due to the questionnaire not being mandatory for MWMP participants (see **Table 3S**) and results showed a non-significant association of TFEQ Cognitive Restraint scores predicting long-term weight loss during the maintenance period [*R^2^*=0.093, *F*(1,18)=1.84, *p*=0.19, **Figure 4b**, **Table 3S**]. The association between aMCC glutamate modulation and the TFEQ Cognitive Restraint score was significant [*R^2^*=0.20, *F*(1,18)=4.56, *p*=0.047, **Figure 4c**, **Table 3S**] reflecting a lower glutamate modulation in those participants reporting lower TFEQ Cognitive Restraint scores. Though not significant, the statistics with the other two TFEQ sub-scores are shown in **Table 3S**.

**Figure 4.**
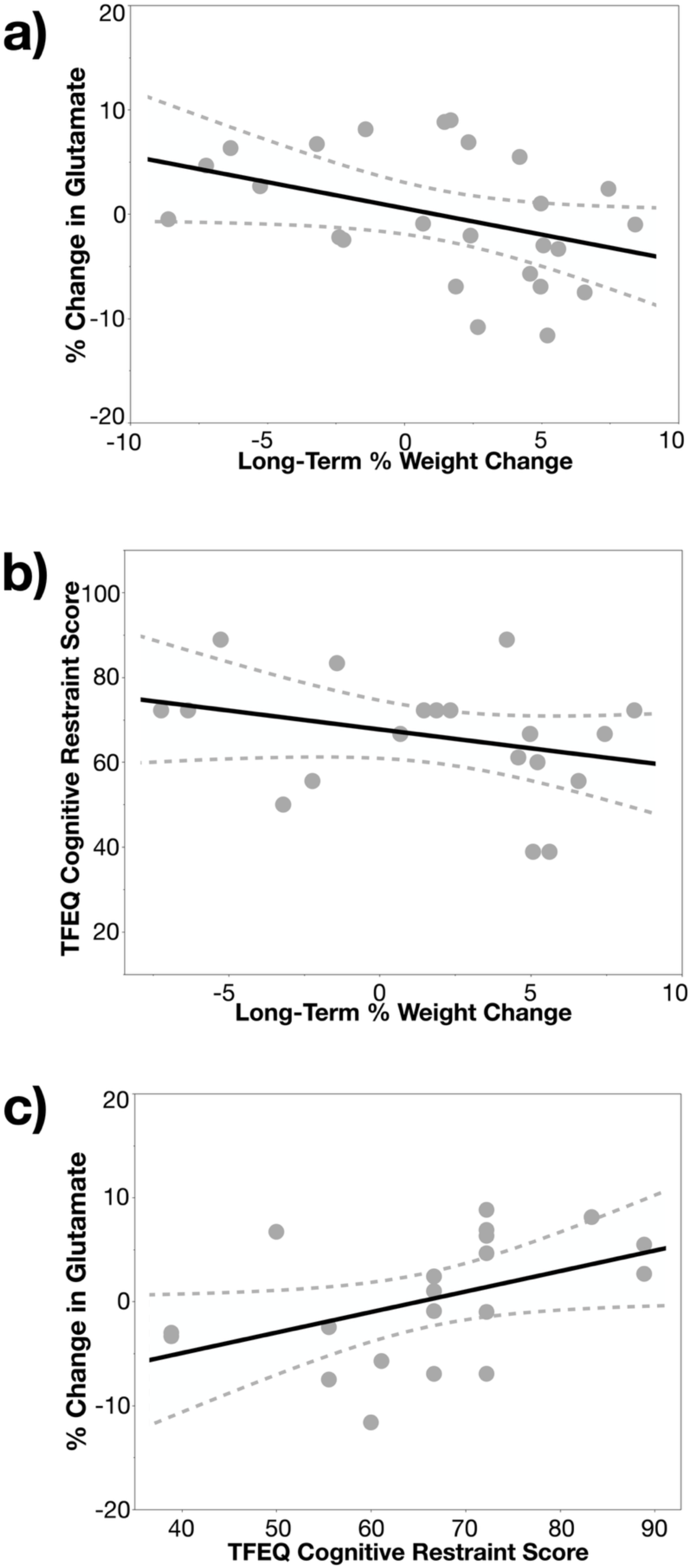
Associations between aMCC glutamate modulation, long-term weight loss performance and self-reported eating behaviors. a) Depicts a trending association between aMCC glutamate modulation during the selective responding mode with food/non-food cues and percent change in weight during the weight maintenance period of the MWMP. b) A non-significant correlation between the TFEQ Cognitive Restraint score and the percent change in weight during the weight maintenance period. c) A significant correlation between aMCC glutamate modulation during the selective responding mode with food/non-food cues and the TFEQ Cognitive Restraint score. See text for the statistical results.

## Discussion

Poor inhibitory control has been implicated in obesity, and it has been observed that these inhibitory control difficulties are likely specific to the presence of food-related cues, i.e., the performance of top-down cognitive control being “distracted” by appetitive food cues eliciting urges to eat [4,7,8]. However, the neurobiology is poorly understood. This study leveraged a unique sample from the MWMP and an innovative task-based ¹H fMRS approach to investigate the impact of food cues on the neurobiological response of glutamate from the aMCC during response inhibition. Qualitatively, aMCC glutamate modulation driven by task demand utilizing neutral (non-affect) cues tended to be positive (relative to the baseline control condition; **Figure 3**). However, switching the task cues to appetitive food cues drove a significantly lower glutamate modulation during the responding mode requiring response inhibition (**Figure 3**). Participants also demonstrated a behavioral stimulus type effect with poorer false alarm rates with food/non-food cues (vs neutral cues). More importantly, participants with OV/OB who tended to show poorer ability in maintaining long-term weight loss post-intervention and poorer TFEQ Cognitive Restraint scores also demonstrated a greater impact (or reduction) on the aMCC glutamate modulation (selective mode with food/non-food cues). Together, these highly novel results demonstrate an effect of aMCC glutamate modulation being impacted by the presence of appetitive food cues compared to neutral cues in individuals with OV/OB. The degree of this (reduced) modulation has a relationship with self-reported eating behaviors and fell short of being related to long-term weight loss maintenance.

The magnitude of the glutamate modulation from the aMCC would be expected not to differ regardless of target cue type (neutral vs appetitive food/non-food images) in individuals less distracted by appetitive food cues (good top-down control). In the presence of neutral cues, the aMCC glutamate modulated tended to be positive during both responding modes with group means (and 95% CI) of 1.71% (-0.46–3.88%) for non-selective and 2.61% (0.48–4.74%) for selective. The glutamate modulation is relative to the glutamate level acquired during a visual flashing checkerboard condition, the baseline control condition that does not require any inhibitory or motor control processes. Increased glutamate modulation therefore suggests a net increase in aMCC excitatory neuromodulation drive [20–23]. However, the aMCC glutamate modulation during processes requiring response inhibition was impacted by the presence of food/non-food cues, as demonstrated by a significantly lower glutamate modulation. This decreased modulation would suggest a decreased excitatory neuromodulation drive in the aMCC, consistent with the framework of appetitive food cues acting as a distractor on top-down inhibitory control processes due to competing resources from bottom-up emotional centers attuned to the food cues [12,39–41]. Future studies are needed to directly investigate potential impact of bottom-up neural processes on top-down inhibitory control with food cues in obesity.

The significant association with self-reported confidence in suppressing urges to eat suggests a relationship between increased top-down inhibitory control neurobiology in the presence of food cues and increased inhibitory control behaviors which would support successful weight loss maintenance. The trending correlation between glutamate modulation and weight loss maintenance suggests there may be a relationship between underlying top-down inhibitory control neurobiology and the ability to maintain weight loss. However, this trend should be viewed with caution. This finding is potentially consistent with previous work using BOLD fMRI, which has identified an association between increased activity in top-down inhibitory control regions, including work assessing the aMCC, and greater success in weight loss and weight loss maintenance [14,39,41–44]. However, as these results did not achieve significance, this the connection between decreased top-down inhibitory control excitatory drive in the presence of food cues and weight changes still warrants further study.

The lack of correlation between aMCC glutamate modulation and task performance was consistent with previous findings [36]. More importantly, the significant stimulus type effect for false alarm rate is consistent with previous meta-analysis findings that food cues drove a significantly poorer false alarm rate in individuals with obesity [7]. The consistency in these findings highlights the importance of utilizing food cues as a distractor in assessing response inhibition in individuals with OV/OB.

In summary, decreased aMCC glutamate modulation was observed in the presence of appetitive food cues compared to neutral squares cues during response inhibition in individuals with OV/OB attempting to maintain weight loss. The relationship between increased aMCC glutamate modulation during selective inhibitory motor responding in the presence of food cues and higher confidence in suppressing urges to eat suggests a connection between underlying neurobiology and inhibitory control behaviors that may promote successful weight loss maintenance. Future ^1^H fMRS research is needed to confirm the trending relationship of weight loss maintenance with neurobiology and assess food-cue driven neurobiology changes in bottom-up emotional centers during inhibitory control.

## Data Availability

Data are available from the corresponding author with reasonable request.

## Acknowledgements

The authors would like to acknowledge and thank Dr. John France for his assistance in setting up the task presentation for this study. Publicly available tools were used for data analysis (LCModel and SAS). Data are available from the corresponding author with reasonable request.

## Funding

This work was supported by pilot funds from the Michigan Diabetes Research Center (UM SUBK00014729 MDRC).

## Disclosure

The authors declare no conflict of interest.

## Supplemental Material

**Supplemental Table 1.**
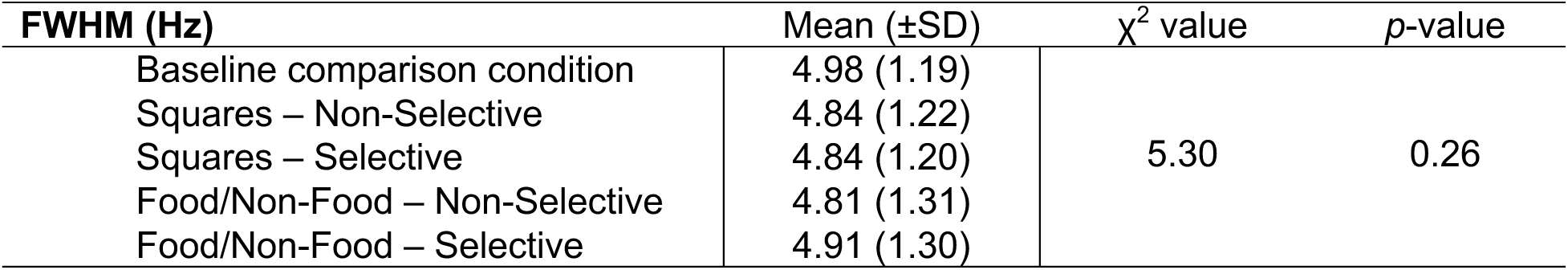
Mean full-width-at-half-max in Hz during the baseline control condition (fixation checkerboard) compared to the task conditions.

**Supplemental Table 2.** Model terms assessing motor response latency, response hit rate, and false alarm rate, including sex and age as covariates. The sex term was significant for response hit rate (*χ^2^*=4.29, *p*=0.038) demonstrated significantly greater errors in females compared to males (means±SEM; 72.8±2.7% v. 94.0±1.4%).

| Response Latency Model Terms | Non-Selective |  | Selective |  |
| --- | --- | --- | --- | --- |
| | $\chi^2$ value | *p*-value | $\chi^2$ value | *p*-value |
| aMCC glutamate modulation | 1.28 | 0.26 | 2.17 | 0.14 |
| Stimulus type (squares or food/non-food) | 0.47 | 0.49 | 2.84 | 0.09 |
| Sex | 0.16 | 0.69 | 0.99 | 0.32 |
| Age | 0.87 | 0.35 | 0.30 | 0.59 |
| Response Hit Rate Model Terms | $\chi^2$ value | *p*-value | $\chi^2$ value | *p*-value |
| aMCC glutamate modulation | 0.02 | 0.89 | 0.79 | 0.37 |
| Stimulus type (squares or food/non-food) | 0.01 | 0.92 | 3.34 | 0.07 |
| Sex | 3.05 | 0.08 | 2.76 | 0.10 |
| Age | 0.25 | 0.62 | 0.07 | 0.79 |
| False Alarm Rate Model Terms | | | $\chi^2$ value | *p*-value |
| aMCC glutamate modulation |  |  | 0.03 | 0.86 |
| Stimulus type (squares or food/non-food) |  |  | 5.14 | **0.023** |
| glutamate modulation by Stimulus type Interaction |  |  | 2.57 | 0.11 |
| Sex |  |  | 0.32 | 0.57 |
| Age |  |  | 0.37 | 0.54 |

**Supplemental Figure 1.**
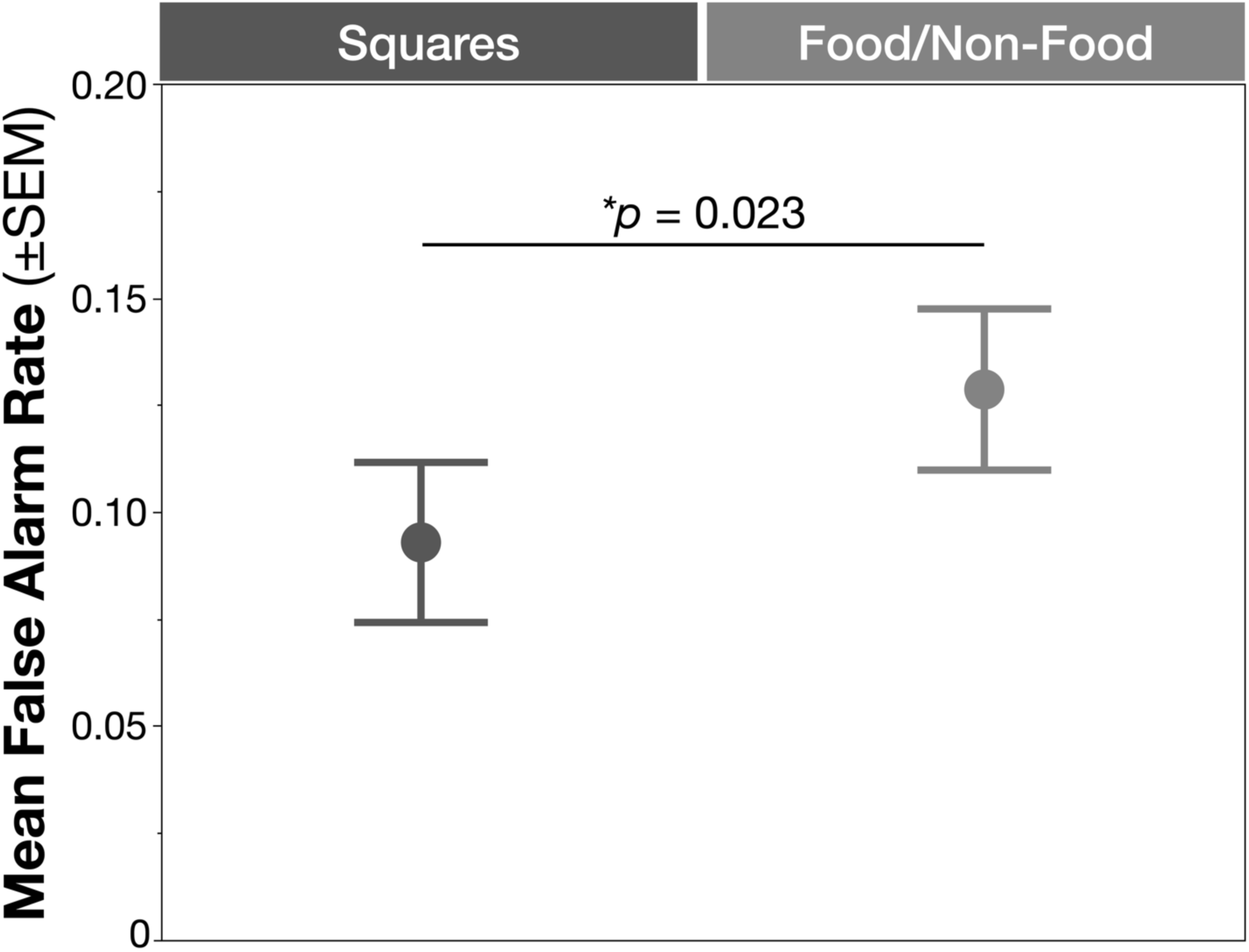
Mean false alarm rate. The stimulus type term was significant (χ^2^ =5.14, p=0.023), supporting a higher false alarm rate with Food/Non-Food stimuli compared to neutral Squares stimuli.

**Supplemental Table 3.** Mean TFEQ values at six-months after beginning the MWMP. Long-term Weight loss performance and self-report ratings on eating behavior with correlations.

| Table 3S: Long-term Weight loss performance and self-report ratings on eating behavior with correlations. |  |  |  |
| --- | --- | --- | --- |
| Three-Factor Eating Questionnaire Scores (6-month) | Mean (±SD) | Range |  |
| Cognitive Restraint | 68.1 (14.8) | 38.9 to 94.4 |  |
| Emotional Eating | 36.9 (24.6) | 0.0 to 66.7 |  |
| Uncontrolled Eating | 39.4 (24.0) | 3.7 to 81.5 |  |
| TFEQ Correlations | $R^2$ | $F$ (DF) | $p$-value |
| Cognitive Restraint |  |  |  |
| % Change in Glutamate | 0.20 | 4.56 (1,18) | 0.047 |
| **Long-Term % Weight Change** | 0.093 | 1.84 (1,18) | 0.19 |
| **Emotional Eating** |  |  |  |
| **% Change in Glutamate** | 0.003 | 0.05 (1,18) | 0.82 |
| **Long-Term % Weight Change** | 0.002 | 0.04 (1,18) | 0.84 |
| **Uncontrolled Eating** |  |  |  |
| **% Change in Glutamate** | 0.012 | 0.22 (1,18) | 0.65 |
| **Long-Term % Weight Change** | 0.015 | 0.27 (1,18) | 0.61 |

**Supplemental Table 4.** MRS reporting parameters, in accordance with “Minimum Reporting Standards for in vivo Magnetic Resonance Spectroscopy (MRSinMRS): Experts’ consensus recommendations,” Lin et al., 2021

|  |  |
| --- | --- |
| **1. Hardware** |  |
| a. Field Strength | 3 T |
| b. Manufacturer | Siemens |
| c. Model (software version if available) | Verio (VB17) or Cima.X (XA61 A-sp02) |
| d. RF coils: nuclei (transmit/receive), number of channels, type, body part | 32 channel volume head coil |
| e. Additional hardware | N/A |
| **2. Acquisition** |  |
| a. Pulse sequence | PRESS |
| b. Volume of interest (VOI) locations | Dorsal anterior cingulate cortex |
| c. Nominal VOI size [cm<sup>3</sup>, mm<sup>3</sup>] | 20 x 17 x 12 mm<sup>3</sup> |
| d. Repetition time ($T_R$), echo time ($T_E$) [ms, s] | *Water Suppressed:* $T_R$= 4-4000ms, $T_E$= 23ms<br>*Water Unsuppressed:* $T_R$= 10sec, $T_E$= 23ms |
| e. Total number of excitations or acquisitions per spectrum | 16 measurements/scan |
| In time series for kinetic studies | i. 4 averages |
| i. Number of averaged spectra (NA) per time point | ii. Block-wise |
| ii. Averaging method (eg block-wise or moving average) | iii. 4 spectra |
| iii. Total number of spectra (acquired/in time series) |  |
| f. Additional sequence parameters (spectral width in Hz, number of spectral points, frequency offsets) | 2000 Hz; 2048 points |
| If STEAM: mixing time ($T_M$) | |
| If MRSI: 2D or 3D, FOV in all directions, matrix size, acceleration factors, sampling method |  |
| g. Water suppression method | VAPOR |
| h. Shimming method, reference peak, and thresholds for “acceptance of shim” chosen | FASTESTMAP |
| i. Triggering or motion correction method (respiratory, peripheral, cardiac triggering, incl. device used and delays) | N/A |
| **3. Data analysis methods and outputs** |  |
| a. Analysis software | LCModel |
| b. Processing steps deviating from quoted reference or product | Adjustment to handling of 1<sup>st</sup> order phase and glutamate/glutamine ratio soft constraint (see Eichstaedt et al., 2026) |
| c. Output measure (eg. absolute concentration, institutional units, ratio) | Absolute levels and expressed as institutional units |
| d. Quantification references and assumptions, fitting model assumptions | Fully relaxed unsuppressed water signal; corrected for differences in volume between water suppressed and unsuppressed (CSF tissue fraction), T<sub>1</sub> and T<sub>2</sub> relaxation on the metabolites, and adjusted the [H<sub>2</sub>O] based on tissue fraction (GM plus WM only and not CSF) and T<sub>1</sub> and T<sub>2</sub> relaxation on the tissue fractions; simulated basis set |
| **4. Data quality** |  |
| a. Reported variables (SNR, linewidth (with reference peaks)) | SNR (mean±SEM): 7.8±0.1; FWHM (mean±SEM): 4.9±0.1 |
| b. Data exclusion criteria | CRLB > 20%; scan CV > 15; SNR < 5 |
| c. Quality measures of postprocessing model fitting (eg CRLB, goodness of fit, SD of residual) | CRLB of Glu (mean±SD): 8.0±1.4 |
| d. Sample spectrum | Figure 2 |

## Notes

### Competing Interest Statement

The authors have declared no competing interest.

### Author Declarations

The Institutional Review Board at Wayne State University gave ethical approval for this work (IRB-21-01-3088).

